# 5hmC-profiles in Puerto Rican Hispanic/Latino men with aggressive prostate cancer

**DOI:** 10.1101/2024.10.26.24315621

**Authors:** Manishkumar S Patel, Mousa Almubarak, Jaime Matta, Carmen Ortiz-Sanchez, Jarline Encarnacion, Gilberto Ruiz-Deya, Julie Dutil, Jasreman Dhillon, Kosj Yamoah, Anders Berglund, Hyun Park, Deepak Kilari, Yoganand Balagurunathan, Liang Wang, Jong Y. Park

**Affiliations:** Department of Tumor Microenvironment and Metastasis, H. Lee Moffitt Cancer Center and Research Institute, Tampa, FL 33612, USA; Department of Basic Sciences, Ponce Research Institute, Ponce Health Sciences University-School of Medicine, Ponce, Puerto Rico; Department of Pathology, H. Lee Moffitt Cancer Center and Research Institute, Tampa, FL 33612, USA; Department of Radiation Oncology, H. Lee Moffitt Cancer Center and Research Institute, Tampa, FL 33612, USA; Department of Biostatistics and Bioinformatics, H. Lee Moffitt Cancer Center and Research Institute, Tampa, FL 33612, USA; Department of Cancer Epidemiology, H. Lee Moffitt Cancer Center and Research Institute, Tampa, FL 33612, USA; Division of Oncology, Medical College of Wisconsin, Milwaukee, WI 53226, USA; Department of Machine Learning, H. Lee Moffitt Cancer Center and Research Institute, Tampa, FL 33612, USA

**Keywords:** 5hmC, Prostate cancer, DNA Methylation, Aggressive Cancer, Puerto Rican Hispanic/Latino

## Abstract

Puerto Rican (PR) Hispanic/Latino (H/L) men are an understudied population that has the highest prostate cancer (PCa) specific mortality among other Hispanic populations. Little information is known about the higher mortality in PR H/L men. It is thought that epigenetic changes in key genes may play a critical role in aggressive tumors. We aimed to identify key 5-hydroxymethylcytosine (5hmC) changes in PR H/L men with aggressive PCa. We performed sequencing analysis using the 5hmC-enriched DNA from 22 prostate tumors and 24 adjacent normal FFPE samples. We identified 808 differentially methylated genes (DMGs) in tumors compared to adjacent normal tissues (FDR<0.05, log2FC>|0.4|). Pathway analysis of DMGs demonstrated that DNA repair pathway was most upregulated in tumors. Since 5hmC abundance positively correlates with gene expression levels, we further investigated 808 DMGs in TCGA PCa gene expression data. Further, we identified 59 DMGs (80.1%, FDR<0.05, ΔGE (gene expression) >|1|) with significant gene expression changes in the same direction. Additionally, we identified 111 aggressiveness-related DMGs, of which, two hypomethylated genes (*CCDC122*, *NUDT15*) and four hypermethylated genes (*PVT1*, *RPL30*, *TRMT12*, *UBR5*) were found to be altered at transcriptomic level in a concordant manner in PR H/L PCa patients (N=86). The aberrant 5hmC (N=55) and GE (N=497) changes in these six genes were also associated with progression-free survival in the mixed PCa population. In conclusion, our study identified 59 DMGs showing concordant epigenetic and transcriptomic changes in tumor tissues and 111 DMGs showing association with aggressive PCa among PR H/L men.

## Introduction

In 2024, 299,010 new prostate cancer (PCa) cases and 35,250 PCa-specific deaths are anticipated in the US (Siegel et al., 2024). The lifetime risk of PCa in US men is approximately 12.5% (NCI, 2018). PCa-specific mortality (PCSM) rates have been found to vary among different racial/ethnic groups in the US, especially in certain Hispanic subgroups as compared to non-Hispanic Whites (NHWs) and non-Hispanic Blacks (NHBs) (Chinea et al., 2017). This study combined all Hispanic subgroups into one broad group including Mexican Americans, Caribbeans, Puerto Ricans, and South Americans. However, among different Hispanic/Latino (H/L) subgroups, Puerto Rican men showed significantly higher PCSM rates than other Hispanic groups and NHBs (Chinea *et al*., 2017). Indeed, PCa is the most common cancer case and cancer-specific death in Puerto Rico (Tortolero-Luna, 2013). According to the 2019 PR Cancer Registry data, PCa is the leading cancer type in terms of incidence (3,528/8,782, 40% of all male cancer cases) and mortality (406/2,703, 15% of all male cancer deaths) in Puerto Rican (PR) H/L men. Despite the high level of vulnerability in this population, the underlying causes of the high mortality rate in this group are still unclear.

PCa is a complex disease that is mediated by the accumulation of genetic and epigenetic aberrations, such as differential expression of oncogenes and tumor suppressor genes (Mateo et al., 2015). Differential DNA methylation can influence carcinogenesis and disease progression (Xu et al., 2019). Epigenetic changes such as DNA methylation (5hmC and 5mC) are an important mechanism responsible for transcription regulation and ultimately functional implications to drive aggressive pathology of PCa (Sjostrom et al., 2022; Zhao et al., 2020). Zhao et al. reported that differential 5mC changes during PCa progression at putative regulatory regions. Indeed, the most common molecular events in PCa is DNA methylation dysregulation. Among these epigenetic changes, some specific changes may be associated with poor outcomes, including PCSM, metastasis, and recurrence (Massie et al., 2017). The Cancer Genome Atlas (TCGA) study found associations between gene expression and methylation profiles. This study suggested that epigenetic changes define distinct molecular subtypes of PCa (Cancer Genome Atlas Research, 2015). The role of DNA methylation in promoter regions has been investigated numerously, and many differentially methylated genes have been related to gene silencing of tumor suppressor genes in PCa and with poor outcomes (Carleton et al., 2018; Giudice et al., 2017; Komura et al., 2018; Xu *et al*., 2019).

In addition to commonly known 5-methylcytosine methylation in the genome, 5-hydroxymethylcytosines (5hmC) are also reported. These 5hmCs are created by oxidation of common 5-methylcytosine methylation by ten-eleven translocation (TET) enzymes (Carter et al., 1992). Several studies reported a regulatory role of 5hmC in gene expression (Mooijman et al., 2016; Sjostrom et al., 2022). Like common 5-methylcytosine methylation, locations of 5hmC are in gene bodies, promoters, and enhancers, which are transcriptionally active regions (Han et al., 2016). Therefore, 5hmCs were suggested as a new class of epigenetic biomarkers for various cancers, including PCa (Nishiyama and Nakanishi, 2021; Sjostrom *et al*., 2022; Xiao et al., 2021).

Notably, 5hmC modification is predominant in gene bodies and is a more accurate marker in echoing gene expression than gene body 5mC (He et al., 2021). Also, around 33% of 5hmC peaks are in tissue-specific differentially methylated regions potentially affecting tissue-specific functional gene expression (Cui et al., 2020; He *et al*., 2021). 5hmC DNA methylation also has an essential tissue-specific function in epigenomic activation in PCa; it was identified as a potential biomarker of aggressive PCa (Sjostrom *et al*., 2022). However, it is not known why PR H/L men show high PCa-specific mortality. Since differential DNA methylation may influence racial disparities in PCa, there is a need to investigate 5hmC profiles to evaluate potential PR-specific methylated genes associated with poor prognosis. To identify promising 5hmC biomarkers for aggressive PCa in PR men, we applied the 5hmC-Seal technology (Song et al., 2011) to examine methylation changes in PCa tissues from PR men. Our results suggested that differential 5hmC changes in a group of candidate genes are associated with aggressiveness and potentially contribute to cancer disparity.

## Methods

### IRB Approval and Tissue Sample Selection

Two Institutional Review Boards, the Moffitt Cancer Center (Protocol no. Pro00048100) and the Ponce Health Sciences University (PHSU) (Protocol no. 1909021277A001), approved this study.

All study participants signed an Informed Consent. We obtained 86 formalin-fixed paraffin-embedded (FFPE) prostate tumor and adjacent non-involved pair samples from the Puerto Rico Biobank (PRBB), a U54 PHSU-MCC PACHE Partnership core facility. Based on Gleason scores and following the 2023 National Comprehensive Cancer Network (NCCN) guidelines for prostate cancers, tumors from study participants were classified as either aggressive or indolent.

### Gene expression

mRNA transcript quantification was done using the Human Exon 1.0 ST microarray (Thermo-Fisher, Carlsbad, CA, USA) at the Genomic Core at Moffitt Cancer Center. RNA was extracted from the FFPE blocks using macro-dissection. The microarray measures 46,050 RNA transcripts. The SCAN (Piccolo et al., 2012) algorithm was used to preprocess and normalize the transcriptomic data resulting in log2 gene expression. Decipher, a 22-marker prognostic gene-expression score, was determined from the Decipher Prostate cancer classifier assay (Marrone et al., 2015; Piccolo *et al*., 2012).

### DNA extraction and quality control

Genomic DNA samples were obtained from the FFPE prostate tumor tissues as described in the manufacturer’s instructions (QIAamp DNA FFPE Tissue kit, Qiagen, Germantown, MD). DNA was extracted from the marked tumor area on the H&E slides by the pathologist (J.D.) from 46 tumor tissues from PR H/L patients. DNA quality was tested with DNA integrity numbers (DINs) and quantified using Qubit.

### 5hmC Library preparation

We used 7-50 ng of genomic DNA as starting material. Briefly, DNA polishing (at 37°C for 30 min) and enzymatic fragmentation (at 37°C for 5 min.) were carried out using NGS FFPE DNA polishing kit (KAPA/Roche, USA) and DNA fragmentation kit (KAPA/Roche, USA) as per manufacturer’s instructions. After fragmentation, DNA sample was end-repaired and A-tailed using KAPA/Roche Hyper Prep Kit PCR-Free as per manufacturer’s instructions. End-repaired DNA was ligated with adapters (5 NEBNext Multiplex Oligos, Illumina), processed further for USER enzyme digestion, and purified. After digestion, DNA was enriched by labeling and capturing as described previously (Song *et al*., 2011). The enriched DNA was used for qPCR (4 μl) and library amplification (20 μl). Fold-change was calculated by Δ-Δ Ct formula (2^(–ΔΔCt)^) = (ΔCt Sample) – (ΔCt control). The 5hmC DNA was amplified using universal primer (New England Biolabs, USA), index primer (New England Biolabs, USA) and HiFi HotStart ReadyMix (KAPA/Roche). Further, purified libraries were quantified using the Quantus instrument from (Promega) and quality checked using Tapestation. Next, Single-end 75 bp sequencing was performed on an Illumina NextSeq 500. A total of 22 prostate tumors and 24 adjacent normal FFPE samples were sequenced after 5hmC enrichment.

### Sequencing data processing

FastQC was used to evaluate raw read quality (Andrews, 2010). Reads were aligned to human genome build hg38 from Ensembl (https://ftp.ensembl.org/pub/release-111/fasta/homo_sapiens/dna/Homo_sapiens.GRCh38.dna.primary_assembly.fa.gz) using bowtie2 v2.5.1 (Langmead and Salzberg, 2012) and sorted and indexed using samtools v1.17 (Li et al., 2009). Further, duplicates were removed from mapped reads using Picard (Picard, 2019), and, raw read counts per gene were generated using the feature Counts tool from package subread (Liao et al., 2014). Samples were checked for outliers using Principal Component Analysis (PCA).

### Differentially methylated genes (DMGs) and pathway analysis

Differentially methylated genes were identified using the DESeq2 package (Love et al., 2014). All samples were normalized using Deseq2 internal normalization and further compared in unpaired manner with multiple-hypothesis testing. Pathway analysis was performed using GSEA (Subramanian et al., 2005). Top pathways were selected based on p_adj_<0.05. An enhanced Volcano package was used to prepare volcano plot. The cluster Profiler package (Wu et al., 2021) performed pathway analysis and visualized functional profiles of differentially methylated genes.

### Differentially Expressed Genes analysis-TCGA

IlluminaHiSeq pancan normalized prostate cancer gene expression (n=550) data was downloaded from TCGA Hub. From total 550 samples, 52 samples were normal solid tissue biopsies and 497 were primary tumors. To identify differentially expressed genes, we calculated ΔGE (Δ gene expression-differences in GE between tumor and adjacent normal samples) and filtered for genes having ≥ 1 or ≤ −1 value for ΔGE and adjusted p-value<0.05.

### Integration with 5hmC and gene expression changes in PCa patients from mixed origin

5hmC DNA methylation and gene expression values were intergrated for DMGs and differentially expressed genes. Log2FC was used for 5hmC and ΔGE values were utilized for TCGA gene expression data. Significantly hypo- or hypermethylated genes showing the same direction of alteration as previously published 5hmC data or gene expression were plotted on the same plot using the ggplot2 package in R (Wickham 2016).

### Risk analysis in PR H/L men for 5hmC and Gene expression profile in PR H/L men

Gleason score (GS) was used as a cut-off to identify DMGs in aggressive PCa patients (GS-7 (3+4) or less, Non-aggressive; GS-7 (4+3) and above, Aggressive). For Gene expression profile, we used 21 aggressive and unpaired 65 indolent tumors. Unpaired two-sided Wilcoxon rank sum test from the ‘stats’ package was used to calculate significantly different 5hmC or gene expression changes in aggressive vs indolent tumors. Pheatmap package (Kolde, 2019) was used to create heatmap plots and the ggplot2 package (Wickham, 2016) was used to create boxplots.

### cfDNA 5hmC sequencing and survival analysis in advanced prostate cancer patients

The 5hmC enrichment and sequencing library preparation have been published (Li et al., 2023). In brief, cell-free DNAs (cfDNAs) were extracted from 0.4-1ml of platelets-poor plasma using QIAamp DNA Blood Mini Kit (Qiagen). The cfDNA yield was quantified by a Qubit 2.0 Fluorometer (Life Technology) and stored at −80°C until use. As described above, 5-10ng of cfDNA was used for 5hmC enrichment and library preparation.

### Survival Analysis

Survival analysis was performed for 5hmC data generated from cfDNA (N=55) and gene expression data downloaded from TCGA for prostate cancer patients (N=497). For the association study, clinical data for TCGA dataset was also downloaded from GDC (https://xenabrowser.net). We used Kaplan-Meier survival analysis (lower level = below the median and higher level = above the median) to analyze the association of 5hmC or Gene expression levels with PFS. Association with PFS was done using the ‘survival’ package (Therneau, 2020), and graphical representations were created using the ‘pheatmap’, ‘survminer’ and ‘ggplot2’ packages. P<0.05 was considered significant. All statistical analyses were performed in R (v4.3.1).

### Data download

Raw 5hmC-seq data from 51 localized and 7 adjacent normal prostate samples were obtained from European Genome-Phenome (https://ega-archive.org/datasets/EGAD00001008462; Study ID: EGAS00001004942). Gene expression data for prostate cancer samples were downloaded from TCGA (https://tcga-xena-hub.s3.us-east-1.amazonaws.com/download/TCGA.PRAD.sampleMap%2FHiSeqV2_PANCAN.gz).

## Results

### Demographic and Clinicopathological Characteristics of Study Group

Most PR H/L men (90%) are white, while 9% were identified as black based on self-identified information. The mean age at diagnosis for PR H/L men with PCa was 62.8 years (Table 1). Seventy-six percent of all patients (n = 66) had a low Gleason score (6 or 7 (3 + 4)) and were classified as a low-risk group while 24% of all patients (n = 21) had a high Gleason score (7 (4 + 3) or 8−10) and were classified as a high-risk group. As expected, a significantly different distribution in the clinical stage was detected between the two groups (p=0.039). There were no statistically significant differences between the two groups regarding prostate-specific antigen (PSA) levels. The study workflow was divided into two parts (Figure 1). The first part involves a comparison of tumor tissues with adjacent normal controls in PR H/L men and further integration with previously published 5hmC and gene expression datasets of PCa patients from mixed origin. The second part involves risk analysis in PR H/L men with PCa to discover significantly different methylated genes with concordant transcriptomics signatures associated with PCa aggressiveness in PR H/L men. We also validated the DMGs by testing their 5hmC and gene expression levels association with poor survival in PCa patients of mixed origin.

**Figure 1.**
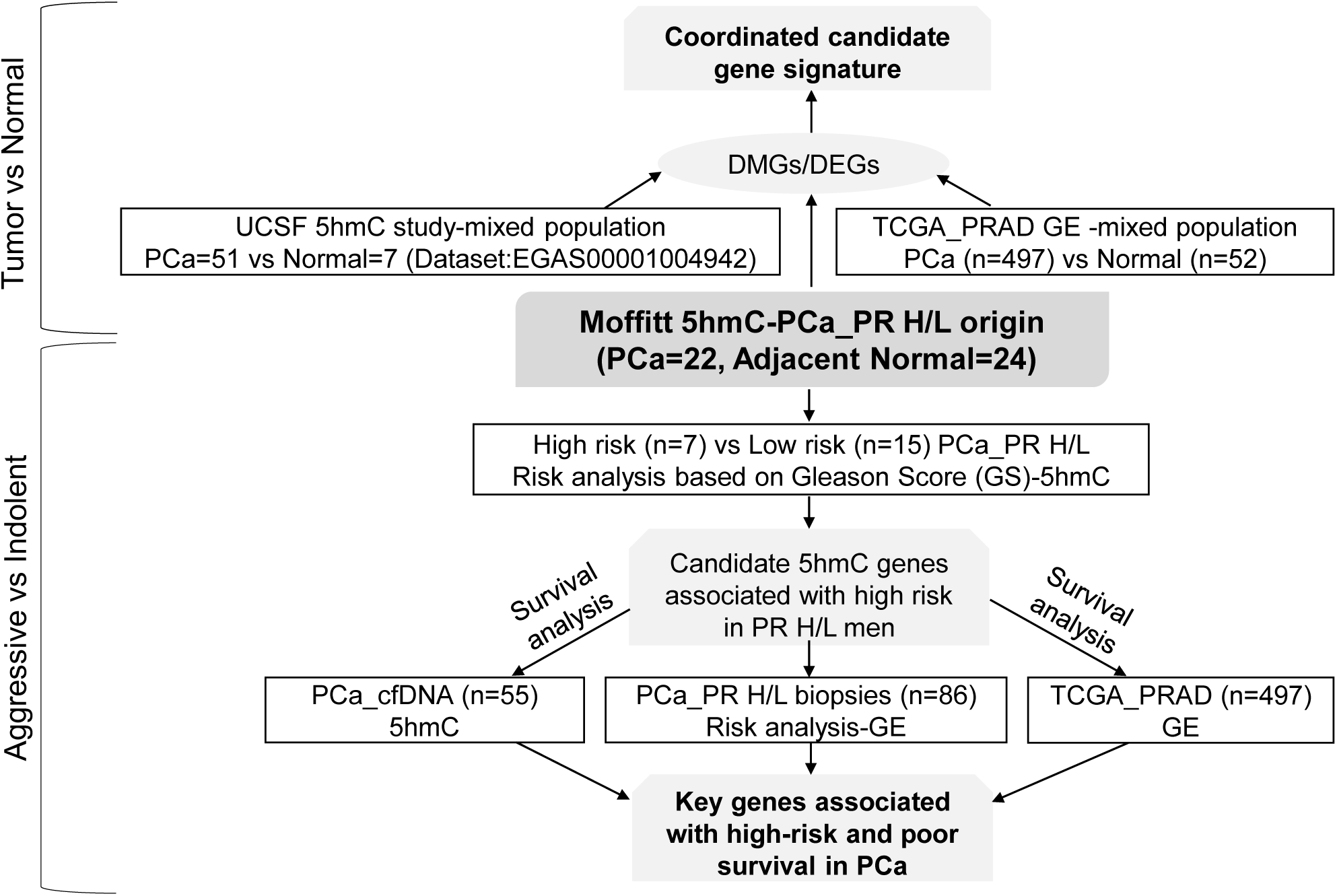
Study workflow. DEGs: Differentially expressed genes; DMGs: Differentially Methylated Genes; GE: Gene Expression.

**Table 1.** Clinicopathological Characteristics of Puerto Rican prostate cancer patients.

| Patients Clinical Characteristics (n=86) |  |  |  |  |  |
| --- | --- | --- | --- | --- | --- |
|  |  | Total (86) | Aggressive (21) | Indolent (65) | pvalue |
| Race | White | 77 | 18 | 59 | 0.22 |
|  | Black | 6 | 2 | 4 |  |
|  | Others | 3 | 2 | 1 |  |
| Stage | T1 or T2 | 69 | 15 | 54 | 0.04 |
|  | T3 | 12 | 6 | 6 |  |
| Gleason score | 6 | 42 | 0 | 42 |  |
|  | 7 (3+4) | 23 | 0 | 23 |  |
|  | 7 (4+3) | 12 | 12 | 0 |  |
|  | 8-9 | 9 | 9 | 0 |  |
| Age |  | 62.8±6.67 | 64.6±5.6 | 61.7±7.1 | 0.41 |
| PSA |  | 6.70±4.3 | 7.82±5.1 | 6.33±4.1 | 0.99 |
| Decipher |  | 0.47±0.19 | 0.50±0.24 | 0.46±0.18 | 0.049 |
| Family history |  | 20.5% | 23.1% | 19.2% | 0.78 |

### Differentially methylated genes in PCa tumors from PR H/L men

To identify differentially methylated genes in PR H/L men, we first compared normalized read counts from 22 tumor samples with 24 adjacent normal samples in an unpaired manner, and identified 808 DMGs (FDR<0.05, log2FC>|0.4|) (Figure 2A, Supplemental Table 1). The most noticeable DMGs included hypermethylated genes (*AGR3, FAM13A, NLRP8, AGAP6, RHPN2, DGAT2L6*) and hypomethylated genes (*IRF2BP1, GPS1, NALT1, HIC1, MAPK7, XKR5, MYBPHL* and *GNAO1*). Since these DMGs may play a key role in PCa, we performed pathway analysis to reveal the biological pathways involved in PCa biology for PR H/L men. This analysis showed that cell cycle, meiosis, cell division, and DNA repair-related pathways were most upregulated in tumor samples (Figure 2B, Supplemental Table 2). This indicates that tumors were highly manipulated with a lack of apoptotic genes and pathways responsible for abnormal growth and survival.

**Figure 2.**
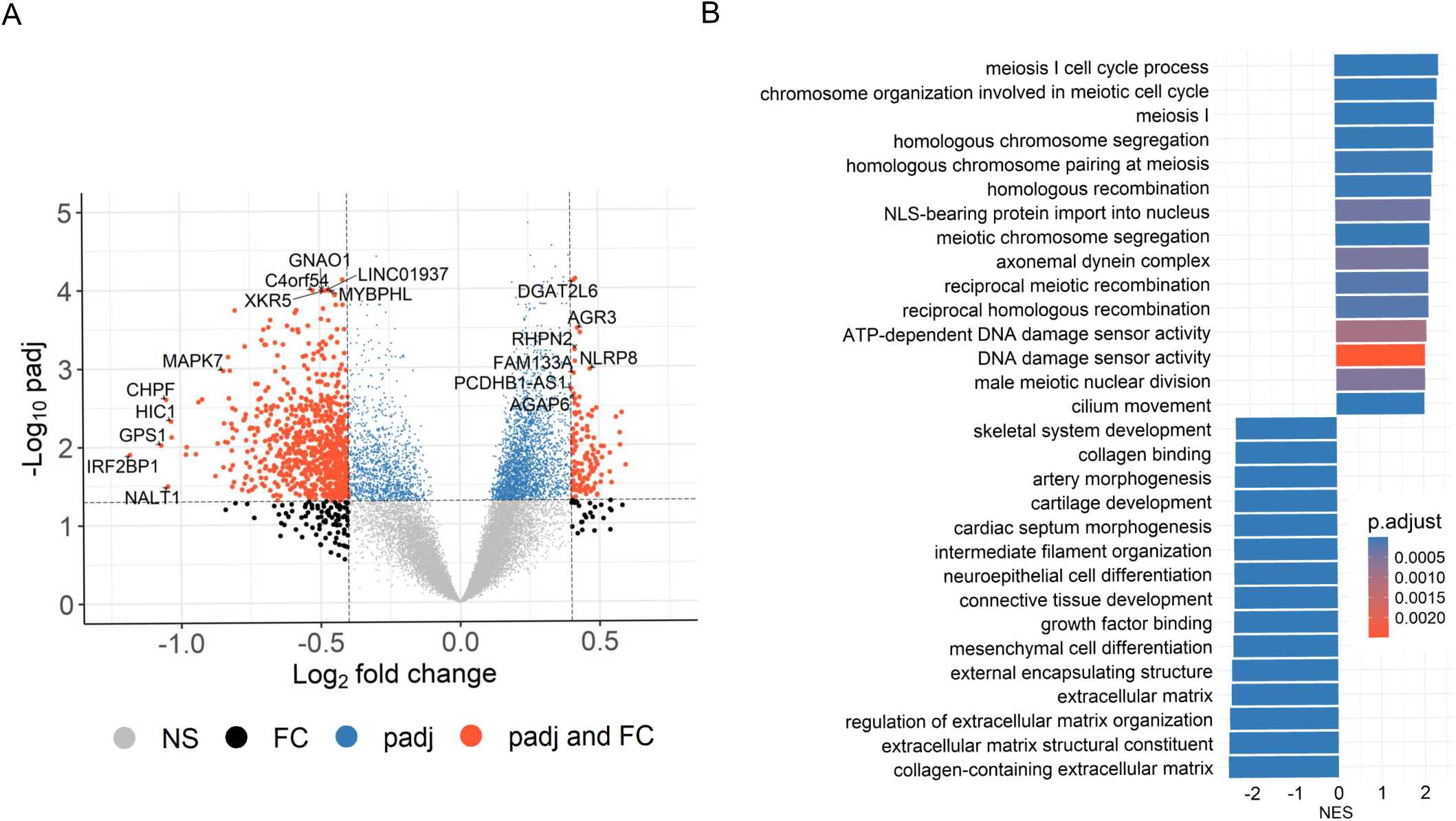
Significant DMGs associated with DNA damage and cell-cycle related pathways in PCa patients of PR H/L origin. (**A**) The volcano plot indicates DMGs in tumor samples (n=22) compared to adjacent normal (n=24); padj<0.05. (**B**) GSEA plot showing top 30 pathways significantly (padj<0.05) altered in tumor tissues compared to adjacent normal tissues. The color intensity represents level of significance.

### 5hmC and gene expression signature in PR H/L men

Since 5hmC abundance is directly correlates with gene expression levels, we investigated 808 DMGs’ expression changes in the TCGA prostate cancer dataset. This analysis identified 59 common DMGs (80.1%, (FDR<0.05, ΔGE>|1|) with significant 5hmC and gene expression changes in the same direction (Figure 3A-3B). This same direction epigenetic and transcriptomic changes (59 genes) in PR H/L men include tumor suppressor genes such as *DKK3* and *PRDM8* with downregulated 5hmC and GE (gene expression) levels (Supplemental Table 3A). We also examined the shared 5hmC candidate genes between PCa patients of the PR H/L population and mixed origin. To identify DMGs in the population with mixed origin, we performed 5hmC methylation analysis in 51 localized PCa and 7 normal samples retrieved from the previously published study dataset (Sjostrom *et al*., 2022). This analysis showed 129 DMGs shared between two populations (Figure 3A, 3C, Supplemental Table 3B). We also observed that the previous 5hmC study showed 171 genes with the same directional gene expression signature in the TCGA dataset. Eight DMGs in PR H/L men have the same direction changes as the other two datasets. Importantly, we found 628 potentially unique 5hmC genes in PR H/L men with PCa (Supplemental Table 3C). These unique differentially methylated genes in PR H/L men include hypomethylated genes such as *IRF2BP1*, *HIC1*, *NALT1*, *MAPK11* and hypermethylated genes such as *CDC25C*, *FLT3, NME5, LDHC* compared to normal samples. Further, we checked whether the 5hmC profile is associated with PCa aggressiveness in PR H/L men.

**Figure 3.**
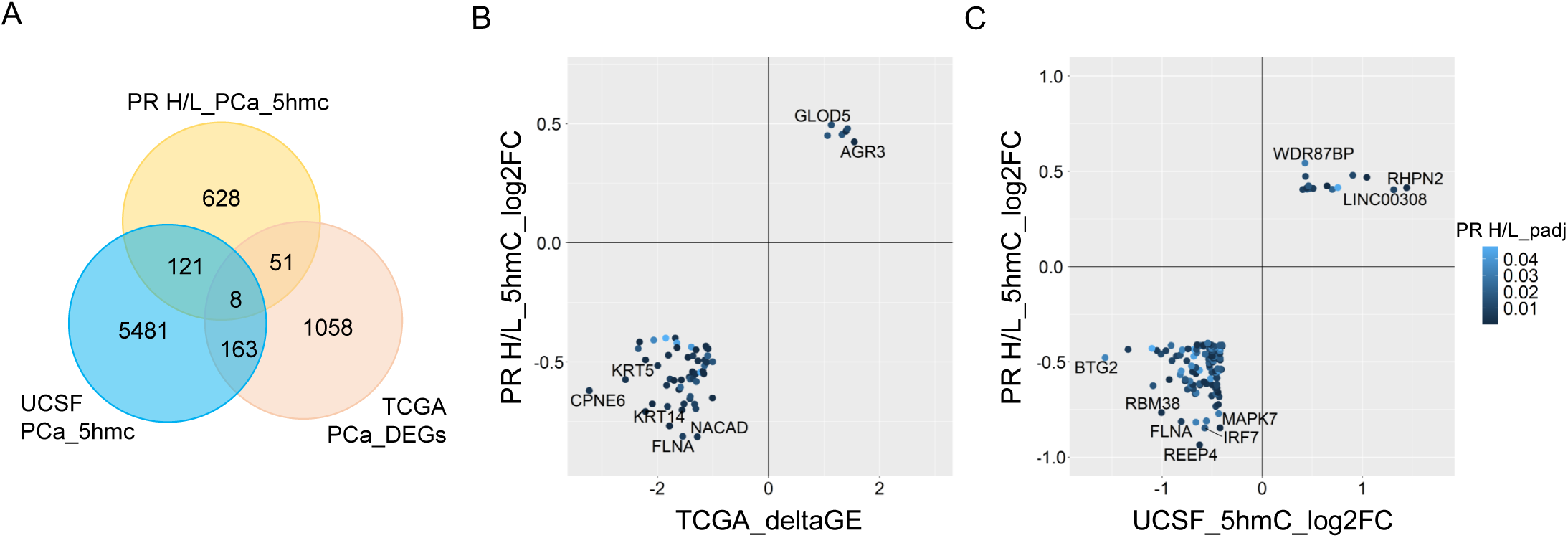
Integration of 5hmC candidate genes from PR H/L PCa patients with UCSF 5hmC and GE dataset from PCa patients of mixed origin. (**A**) Van diagram showing overlapping candidate genes with UCSF 5hmC and TCGA GE datasets (padj<0.05). (**B**) GE (TCGA PCa dataset) and (**C**) 5hmC (UCSF study on PCa patients) changes in the same direction as PR H/L men.

### 5hmC-gene signatures in aggressive tumors

Based on the Gleason score, we classified as 7 aggressive tumors and 15 Indolent tumors in 22 tumor samples from PR H/L men. To detect DMGs associated with high-risk aggressive tumors, we evaluated the difference in 5hmC levels between the two groups and identified 111 DMGs (p<0.05) (Figure 4A,). Among those DMGs, the most noticeable genes included hypomethylated genes (*CCDC122, NUDT15, BCCIP,* and *KLK10*) and hypermethylated genes (*PVT1, TRMT12, RPL30, UBR5*, *COX6C, ARMC2*) in aggressive PCa patients (Supplemental Table 4A). These genes were previously reported for their role in aggressive PCa biology. To check the functional implication of these 111 DMGs, we examined whether their 5hmC levels were correlated with their transcriptomic levels. Out of these 111 DMGs, we confirmed 5hmC hypomethylated genes (*CCDC122* and *NUDT15*) and hypermethylated genes (*TRMT12*, *PVT1*, *RPL30*, and *UBR5*) with same direction GE levels in PR H/L PCa patients (n=86) (Figure 4B-4C, Supplemental Table 4A-4B). These candidate genes in aggressive tumors reveal their significance as potential biomarkers or targets in aggressive PCa patients of PR H/L origin.

**Figure 4.**
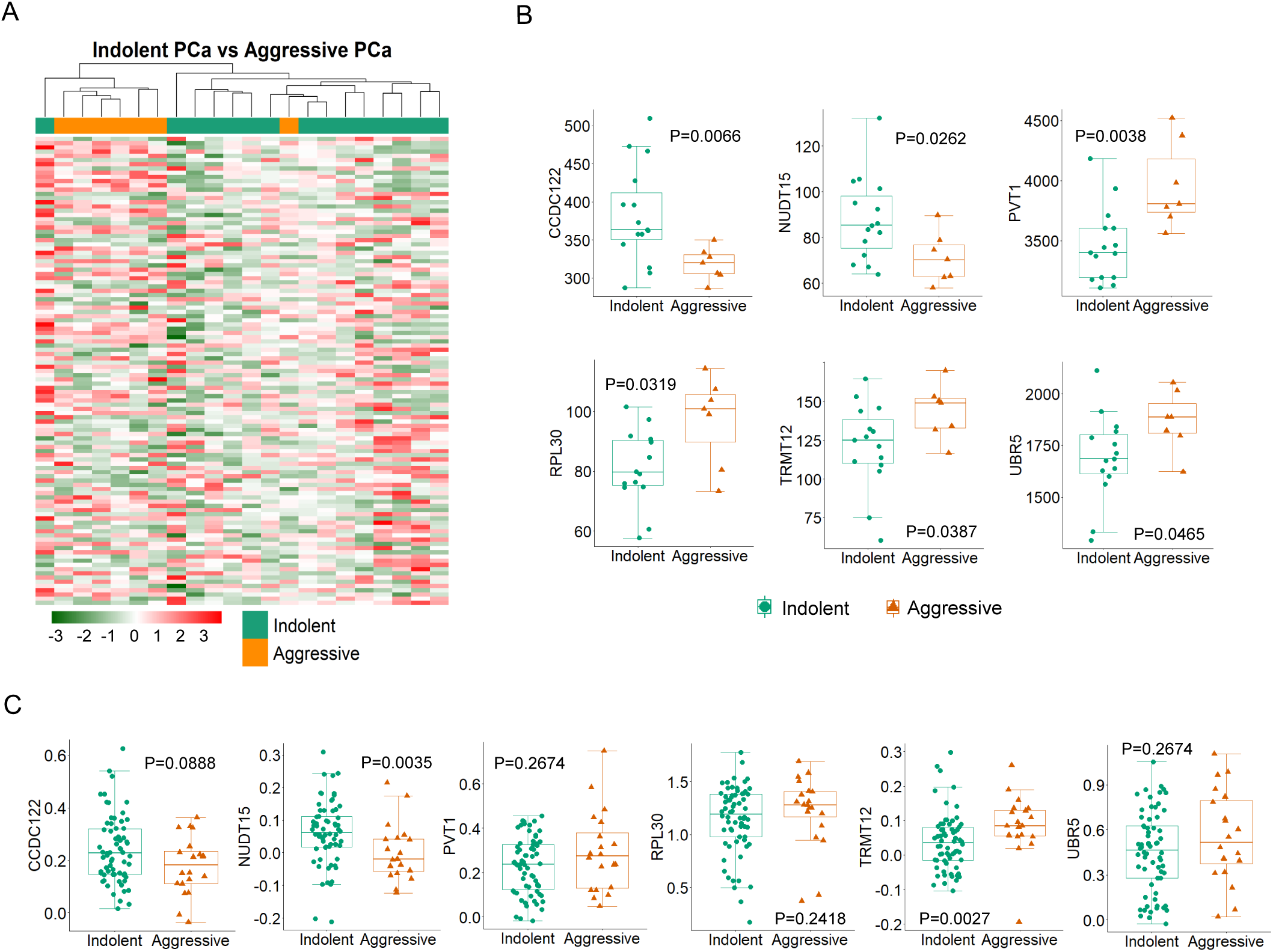
High or low-risk PR H/L PCa patients demonstrated significantly different (P<0.05) and concordant 5hmC-GE signatures. (**A**) The heat map shows 111 DMGs in aggressive patients compared to indolent cases. Representative examples of genes showing association of (**B**) 5hmC levels (n=22) and (**C**) GE levels (n=86) with aggressiveness in PR H/L PCa patients. GS was used to define the risk category of each case. Low risk, GS=<6 & 3+4; High risk, GS=4+3 & >8.

### Association of 5hmC levels with poor progression-free survival

The concordant 5hmC and gene expression signature in *CCDC122, NUDT15, TRMT12, PVT1, RPL30*, and *UBR5* may be responsible for poor survival in PCa patients. However, we could not gather clinical follow-up survival data for PR H/L PCa patients. Hence, we examined the association of these genes with survival in PCa patients of mixed origin. We have used the 5hmC data generated from cfDNA for another study for survival analysis (Li *et al*., 2023). We found that lower levels of *CCDC122* and *NUDT15* and higher levels of *PVT1*, *RPL30*, *TRMT12,* and *UBR5* were significantly associated or trending towards significance while associated with poor PFS in PCa of mixed origin (Figure 5A, Supplemental Table 5A). The consistent findings across diverse DNA sources (tissue biopsy and cfDNA) from PCa patients affirm the significance of altered methylation levels in these genes as reliable indicators for predicting a worse prognosis.

**Figure 5.**
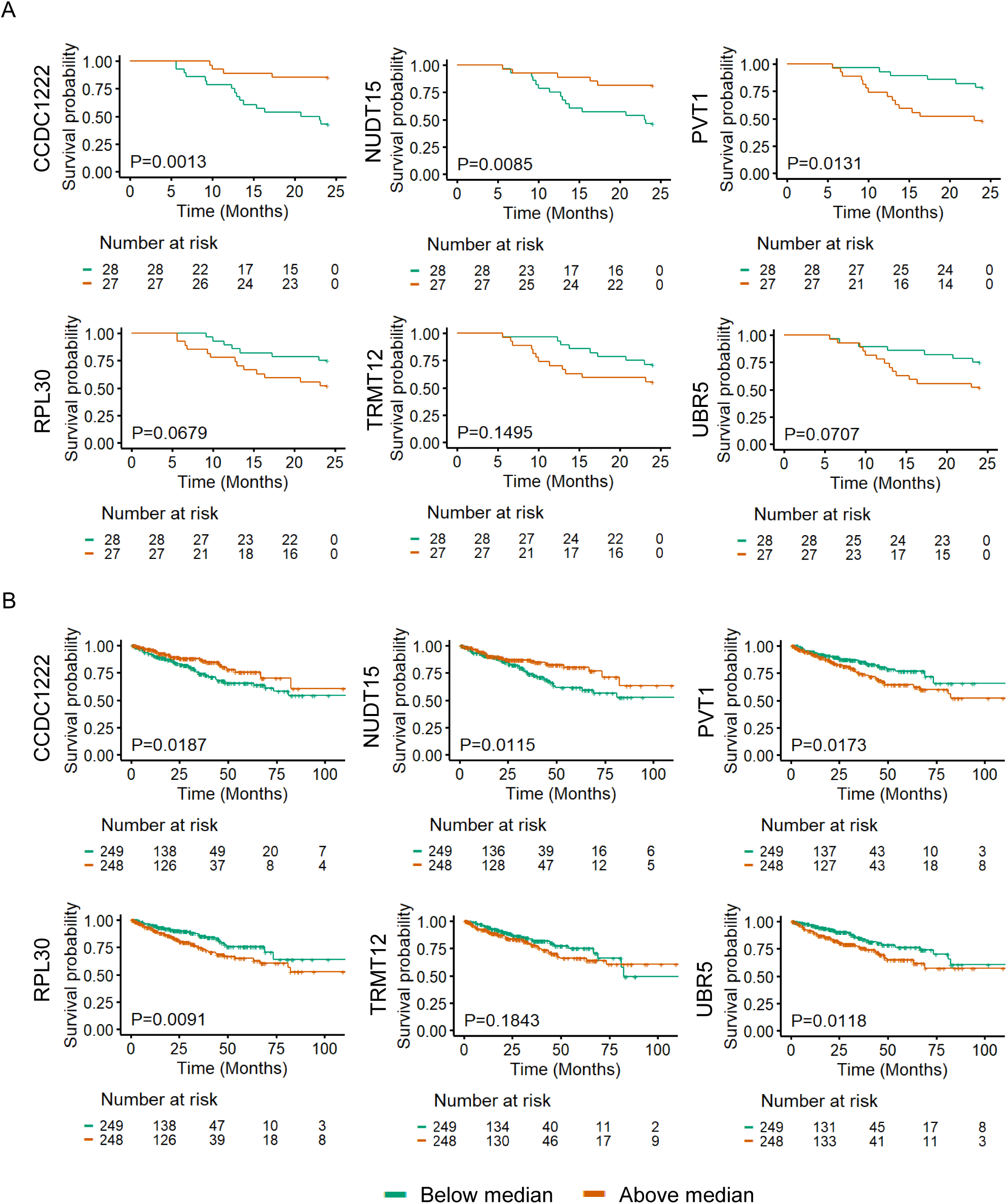
5hmC and GE levels of *CCDC122*, *NUDT15* (low), and *PVT1, TRMT12, RPL30, UBR5* (high) are significantly (P<0.05) associated with poor PFS in PCa patients. Representative examples of genes showing association of (**A**) 5hmC levels in cfDNA (n=55) and (**B**) GE levels in tumor tissues (n=497) with poor PFS in PCa patients of mixed origin. The independent cohorts of PCa patients were used for this analysis. Gene expression levels were retrieved from TCGA database (n=497).

Further, we aimed to investigate whether transcriptomic levels of these genes are also associated with poor PFS in PCa patients. We examined gene expression and survival data from TCGA PCa database (n=497). We found that lower GE levels of *CCDC122* and *NUDT15*, and higher GE levels of *PVT1*, *RPL30, TRMT12*, and *UBR5* showed a clear trend of association with poor prognosis in PCa patients (Figure 5B, Supplemental Table 5B). Our findings revealed a uniform directional cfDNA 5hmC and tissue GE signature of *CCDC122, NUDT15, PVT1, RPL30, TRMT12*, and *URB5*. These genes are associated with poor survival in mixed PCa populations. This further solidifies the role of these candidate genes in aggressive PCa biology.

## Discussion

Despite high PCa-specific mortality, Puerto Rican Hispanic/Latino (PR H/L) men remained an understudied population (Chinea *et al*., 2017). Although PCa is slowly growing, around 20-30% of cases show an aggressive phenotype potentially leading to metastasis and poor survival outcomes. Considering PCa racial and ethnic health disparities, we aim to investigate 5hmC changes in PR H/L PCa patients and their role in the aggressiveness of the disease. Our analysis revealed 59 genes having the same direction of epigenetic and transcriptomic changes in tumor tissue of PR H/L men. Further, we found 111 DMGs associated with PCa aggressiveness with six candidate genes having concordant epigenetic-transcriptomics signatures associated with PCa aggressiveness in PR H/L men. Finally, we demonstrated these candidate genes’ 5hmC and gene expression levels for their association with poor PFS in PCa patients. Our findings provide an essential insight into the epigenetic landscape of PCa in the PR H/L patient population. Some of the genes identified in this study are associated with various cancers including PCa, and affect multiple biological processes, such as immune pathways, cell signaling, metabolism, DNA repair, proliferation, and cell cycle.

Our data indicates significant alterations in the 5hmC profile in PCa tissues compared to normal tissues. The hypermethylated genes include androgen-regulated gene (*AGR3*), and the PCa proliferation-related gene (*RHPN2*). The hypomethylated genes include tumor suppressor genes (*BTG2*, *DKK3* and *PRDM8*), transcriptional repressor (*HIC1, IRF2BP1*), apoptosis related gene (*MAPK7*) and methyltransferases (*PRDM8. PRDM16, PRDM13, KMT5C, FAM86B2, TRMT61A*). AGR3 overexpressed in tumor vs benign and cancer/benign vs castration, meaning that androgen regulate the gene and are potentially involved in prostate carcinogenesis (Vaarala et al., 2012). AGR3 role in AR regulation indicates that it is a candidate target gene. *RHPN2* is a miR-205 target associated with PCa cell proliferation, invasion, and migration (Jiang et al., 2019). *BTG2* is upregulated by *PTEN* and *p53* in human bladder carcinoma cells (Tsui et al., 2018). *DKK3* has a protective role in PCa (Al Shareef et al., 2018). *HIC1* loss promotes PCa metastasis by triggering epithelial-mesenchymal transition (Hao et al., 2017). *IRF2BP1* is a transcriptional corepressor that belongs to the IRF2BP protein family (IRF2BP1, IRF2BP2, and EAP1), and *EAP1* has been reported as a novel AR coregulator (Yokoyama et al., 2021). Overall, we have found upregulated signatures for AR regulation, transcriptional repression, and downregulated tumor suppressor genes. Multiple pathways, such as DNA damage sensor activity, DNA recombination, and cell cycle-related pathways, were upregulated in tumor samples, potentially supporting cancer cell survival, proliferation, and aggressiveness.

The DMGs from our findings also overlapped with a previous 5hmC study (Sjostrom *et al*., 2022) on PCa and gene expression dataset from TCGA. For example, we found *AGR3* and *RHPN2* hypermethylation and *BTG2* and *MAPK7* hypomethylation in both 5hmC datasets. We also found upregulation of *AGR3* and downregulation of *DKK3, PRDM8,* and *TP53AIP1* in our 5hmC study and TCGA gene expression dataset. Interestingly, the PR H/L cohort also showed unique differentially methylated genes, including hypomethylated genes such as *IRF2BP1*, *HIC1*, *MAPK11* and hypermethylated genes such as *CDC25C*, *FLT3*, *NME5, LDHC* (Supplemental Table 3C). These identified gene signatures in PR H/L PCa tumors are highly associated with AR regulation. Downregulation of apoptotic and tumor suppressor pathways leads to prostate cancer aggression, proliferation, survival, and therapeutic resistance.

Previous studies showed that tumor aggressiveness is associated with dysregulation of gene expression in prostate cancer (Ali et al., 2018). We also demonstrated that hypomethylated genes (*CCDC122*, *NUDT15*) and hypermethylated genes (*PVT1*, *RPL30*, *TRMT12*, and *UBR5*) have concordant gene expression changes in PR H/L PCa patients. *CCDC122* and *NUDT15* are located on the nearby cytogenetic band of 13q14.11 and 13q14.2, respectively, and deletion of both genes is associated with PCa growth and survival (Massey et al., 2024; Ongaba, 2023). Notably, allelic loss at 13q14 has been reported in 33% of prostate tumors (Cooney et al., 1996; Yin et al., 1999) and associated with high prostate tumor grade and stage (Dong et al., 2001; Kluth et al., 2018).We believe that hypomethylated 5hmC and downregulated expression of these genes in our study indicate that they could be critical 5hmC markers of PCa in PR H/L men.

We showed that PVT1, *RPL30*, *TRMT12*, and *UBR5* are 5hmC hypermethylated and overexpressed in aggressive tumors. Previous studies showed that these genes are important for AR regulation, PCa growth and aggressiveness. *PVT1* is located on the 8q24 along with c-Myc which well-reported site for copy number gains in different cancers (Graham and Adams, 1986). Previously*, PVT1* promoter 5mC hypomethylation was found to be associated with worse prognosis in renal cell cancer due to *PVT1*-*MYC* upregulation (Posa et al., 2016). *RPL30* (8q22.2), an overexpressed ribosomal proteins (RPs) in PCa (Dolezal et al., 2018), is positively correlated with co-amplification of 8q22-24 regions containing genes encoding the Myc-PVT1 (8q24.21) (Sarver et al., 2016; Tseng and Bagchi, 2015). *TRMT12* (8q24.13), the tRNA methyltransferase, showed strong binding by the AR in castrate-resistant PCa (CRPC) tissue and was overexpressed in CRPC tissue compared to benign or untreated tissue (Sharma et al., 2013). UBR5 (8q22.3) was reported as a top PCa-related E3 ubiquitin ligase which is strongly associated with PC progression and aggressiveness (Yan et al., 2022). It has been demonstrated that chromosome 8q gain is correlated with early progression in hormonal-treated PCa (Steiner et al., 2002). In our study, *PVT1*, *TRMT12*, *RPL30*, and *UBR5* hypermethylation demonstrates that, in addition to 8q gain, 5hmC-based methylation and the corresponding increase in gene expression levels is an important mechanism for higher activity of these genes in PR H/L men with aggressive PCa.

Further, we tested six genes (*CCDC122, NUDT15, PVT1, TRMT12, RPL30*, and *UBR5*) in different cohorts of PCa patients from mixed origin and demonstrated their 5hmC and gene expression levels are associated with poor progression-free survival. Here, the 5hmC levels were measured in cfDNA, which shows that these gene methylation levels can be prominently detected in blood and can be used to predict survival outcomes. The results of this analysis bolster our findings in PR H/L men that these six genes are associated with PCa aggressiveness and, hence, with poor PFS.

The congruent directional changes in 5hmC and gene expression could be critical in aggressive PCa biology; hence, it is worthwhile to validate them as potential biomarkers and therapeutical targets for aggressive PCa among PR H/L men. The novelty of our study is identifying the 5hmC candidate genes and understanding the potential role of 5hmC in an understudied PR H/L population with PCa. As far as we know, this is the first report studying 5hmC methylation in this Puerto Rican population. However, our study has some limitations, which include a small sample size and the unavailability of tumor-normal gene expression data. Due to this, we were not able to directly determine associations with transcriptomic levels of 628 unique DMGs compared to normal tissue in this population. Our ability to perform survival analysis in PR H/L men was also restricted due to the unavailability of clinical follow-up data. In the future, it is important to extend this analysis to a larger cohort with follow-up information and tumor-normal transcriptomics data to identify exclusive genes associated with high specific mortality in PR H/L men with PCa.

In conclusion, our study identifies important gene signatures in RP H/L men with PCa and demonstrates that *CCDC122, NUDT15, PVT1, TRMT12, RPL30*, and *UBR5* are associated with PCa aggressiveness in PR H/L men, hence, poor survival outcomes. The development of biomarkers for PCa aggressiveness will provide more effective tools for the diagnosis of clinically significant disease and facilitate the selection of potential therapeutical drug targets.

## Acknowledgements

This work was supported in part by the U54 Partnership Grant to Address Cancer Health Equity (U54CA163071, MPI: Matta/Dutil and U54CA163068, MPI: Wright/Monteiro) and the Puerto Rico Biobank and Quantitative Sciences Cores. This research was supported by NIH grant R01CA212097 (LW) and R01CA250018 (LW). The results shown in this manuscript are based partly on data generated by the TCGA Research Network: https://www.cancer.gov/tcga. This work has been supported in part by the Molecular Genomics Core at the Moffitt Cancer Center, an NCI-designated Comprehensive Cancer Center (P30-CA076292). The authors also acknowledge the help of David A. Quigley, Ph.D. (Dept. of Urology, UCSF) and The European Genome-phenome Archive (EGA) for providing the access to 5hmC dataset (Study ID: EGAS00001004942).

## Author contributions

M.S.P. and M.A. performed experiments related to the study; J.M., C.O., J.E., G.R., J.Dutil, J. Dhillon, K.Y. provided biopsy tissues; D.K. provided plasma samples; H.P. isolated DNA and RNA; J.M., C.O., J.E., G.R., J.Dutil, J. Dhillon, K.Y., D.K., J.P. collected the clinical information; M.S.P., A.B., J.P., and L.W. analyzed the data; M.S.P., Y.B., J.P., and L.W. wrote the manuscript; J.P., J.M. and L.W. supervised the study and provided funding support.

## Data Availability

The datasets generated and analyzed for this study are available upon reasonable request to the corresponding author.

## Competing Interests

The authors declare no potential conflicts of interest.

## Supplemental Table Legends

**Supplemental Table 1.** 808 differentially methylated genes in tumor samples compared to normal samples from PR H/L men.

**Supplemental Table 2.** Pathway analysis for 808 differentially methylated genes.

**Supplemental Table 3A.** 59 coordinated gene signatures between 5hmC in PR H/L men with PCa and TCGA PCa gene expression dataset.

**Supplemental Table 3B.** 128 coordinated differentially methylated 5hmC genes in PR H/L men with PCa and in mixed population from UCSF 5hmC study on PCa.

**Supplemental Table 3C.** Unique 5hmC gene signature in PR H/L men with PCa

**Supplemental Table 4A.** Differentially methylated genes in aggressive tumors compared to indolent tumors in PR H/L men with PCa

**Supplemental Table 4B**. Association of PCa aggressiveness with gene expression levels of 92/111 differentially methylated genes in PR H/L men with PCa.

**Supplemental Table 5A.** Association of 5hmC levels (in cfDNA) of 94/111 differentially methylated genes with PFS in PCa patients from mixed origin.

**Supplemental Table 5B.** Association of gene expression levels (TCGA PCa dataset) of 82/111 differentially methylated genes with PFS in PCa patients from mixed origin.

## Notes

### Competing Interest Statement

The authors have declared no competing interest.

## References

Al Shareef, Z., Kardooni, H., Murillo-Garzon, V., Domenici, G., Stylianakis, E., Steel, J.H., Rabano, M., Gorrono-Etxebarria, I., Zabalza, I., Vivanco, M.D., et al. (2018). Protective effect of stromal Dickkopf-3 in prostate cancer: opposing roles for TGFBI and ECM-1. Oncogene 37, 5305–5324. 10.1038/s41388-018-0294-0.

Ali, H.E.A., Lung, P.-Y., Sholl, A.B., Gad, S.A., Bustamante, J.J., Ali, H.I., Rhim, J.S., Deep, G., Zhang, J., and Abd Elmageed, Z.Y. (2018). Dysregulated gene expression predicts tumor aggressiveness in African-American prostate cancer patients. Scientific Reports 8, 16335. 10.1038/s41598-018-34637-8.

Andrews, S. (2010). FastQC: A Quality Control Tool for High Throughput Sequence Data [Online].

Cancer Genome Atlas Research, N. (2015). The Molecular Taxonomy of Primary Prostate Cancer. Cell 163, 1011–1025. 10.1016/j.cell.2015.10.025.

Carleton, N.M., Zhu, G., Gorbounov, M., Miller, M.C., Pienta, K.J., Resar, L.M.S., and Veltri, R.W. (2018). PBOV1 as a potential biomarker for more advanced prostate cancer based on protein and digital histomorphometric analysis. Prostate 78, 547–559. 10.1002/pros.23499.

Carter, B.S., Beaty, T.H., Steinberg, G.D., Childs, B., and Walsh, P.C. (1992). Mendelian inheritance of familial prostate cancer. Proc Natl Acad Sci U S A 89, 3367–3371. 10.1073/pnas.89.8.3367.

Chinea, F.M., Patel, V.N., Kwon, D., Lamichhane, N., Lopez, C., Punnen, S., Kobetz, E.N., Abramowitz, M.C., and Pollack, A. (2017). Ethnic heterogeneity and prostate cancer mortality in Hispanic/Latino men: a population-based study. Oncotarget 8, 69709–69721. 10.18632/oncotarget.19068.

Cooney, K.A., Wetzel, J.C., Merajver, S.D., Macoska, J.A., Singleton, T.P., and Wojno, K.J. (1996). Distinct regions of allelic loss on 13q in prostate cancer. Cancer Res 56, 1142–1145.

Cui, X.-L., Nie, J., Ku, J., Dougherty, U., West-Szymanski, D.C., Collin, F., Ellison, C.K., Sieh, L., Ning, Y., Deng, Z., et al. (2020). A human tissue map of 5-hydroxymethylcytosines exhibits tissue specificity through gene and enhancer modulation. Nature Communications 11, 6161. 10.1038/s41467-020-20001-w.

Dolezal, J.M., Dash, A.P., and Prochownik, E.V. (2018). Diagnostic and prognostic implications of ribosomal protein transcript expression patterns in human cancers. BMC Cancer 18, 275. 10.1186/s12885-018-4178-z.

Dong, J.-T., Boyd, J.C., and Frierson Jr., H.F. (2001). Loss of heterozygosity at 13q14 and 13q21 in high grade, high stage prostate cancer. The Prostate 49, 166–171. 10.1002/pros.1131.

Giudice, A., Montella, M., Boccellino, M., Crispo, A., D’Arena, G., Bimonte, S., Facchini, G., Ciliberto, G., Botti, G., Quagliuolo, L., et al. (2017). Epigenetic Changes Induced by Green Tea Catechins a re Associated with Prostate Cancer. Curr Mol Med 17, 405–420. 10.2174/1566524018666171219101937.

Graham, M., and Adams, J.M. (1986). Chromosome 8 breakpoint far 3’ of the c-myc oncogene in a Burkitt’s lymphoma 2;8 variant translocation is equivalent to the murine pvt-1 locus. Embo j 5, 2845–2851. 10.1002/j.1460-2075.1986.tb04578.x.

Han, D., Lu, X., Shih, A.H., Nie, J., You, Q., Xu, M.M., Melnick, A.M., Levine, R.L., and He, C. (2016). A Highly Sensitive and Robust Method for Genome-wide 5hmC Profiling of Rare Cell Populations. Mol Cell 63, 711–719. 10.1016/j.molcel.2016.06.028.

Hao, M., Li, Y., Wang, J., Qin, J., Wang, Y., Ding, Y., Jiang, M., Sun, X., Zu, L., Chang, K., et al. (2017). HIC1 loss promotes prostate cancer metastasis by triggering epithelial-mesenchymal transition. J Pathol 242, 409–420. 10.1002/path.4913.

He, B., Zhang, C., Zhang, X., Fan, Y., Zeng, H., Liu, J., Meng, H., Bai, D., Peng, J., Zhang, Q., et al. (2021). Tissue-specific 5-hydroxymethylcytosine landscape of the human genome. Nat Commun 12, 4249. 10.1038/s41467-021-24425-w.

Jiang, S., Mo, C., Guo, S., Zhuang, J., Huang, B., and Mao, X. (2019). Human bone marrow mesenchymal stem cells-derived microRNA-205-containing exosomes impede the progression of prostate cancer through suppression of RHPN2. J Exp Clin Cancer Res 38, 495. 10.1186/s13046-019-1488-1.

Kluth, M., Scherzai, S., Büschek, F., Fraune, C., Möller, K., Höflmayer, D., Minner, S., Göbel, C., Möller-Koop, C., Hinsch, A., et al. (2018). 13q deletion is linked to an adverse phenotype and poor prognosis in prostate cancer. Genes Chromosomes Cancer 57, 504–512. 10.1002/gcc.22645.

Kolde, R. (2019). Pheatmap: pretty heatmaps. R package version 1, 726.

Komura, K., Sweeney, C.J., Inamoto, T., Ibuki, N., Azuma, H., and Kantoff, P.W. (2018). Current treatment strategies for advanced prostate cancer. Int J Urol 25, 220–231. 10.1111/iju.13512.

Langmead, B., and Salzberg, S.L. (2012). Fast gapped-read alignment with Bowtie 2. Nat Methods 9, 357–359. 10.1038/nmeth.1923.

Li, H., Handsaker, B., Wysoker, A., Fennell, T., Ruan, J., Homer, N., Marth, G., Abecasis, G., Durbin, R., and Genome Project Data Processing, S. (2009). The Sequence Alignment/Map format and SAMtools. Bioinformatics 25, 2078–2079. 10.1093/bioinformatics/btp352.

Li, Q., Huang, C.C., Huang, S., Tian, Y., Huang, J., Bitaraf, A., Dong, X., Nevalanen, M.T., Zhang, J., Manley, B.J., et al. (2023). 5-hydroxymethylcytosine sequencing in plasma cell-free DNA identifies unique epigenomic features in prostate cancer patients resistant to androgen deprivation therapy. medRxiv. 10.1101/2023.10.13.23296758.

Liao, Y., Smyth, G.K., and Shi, W. (2014). featureCounts: an efficient general purpose program for assigning sequence reads to genomic features. Bioinformatics 30, 923–930. 10.1093/bioinformatics/btt656.

Love, M.I., Huber, W., and Anders, S. (2014). Moderated estimation of fold change and dispersion for RNA-seq data with DESeq2. Genome Biol 15, 550. 10.1186/s13059-014-0550-8.

Marrone, M., Potosky, A.L., Penson, D., and Freedman, A.N. (2015). A 22 Gene-expression Assay, Decipher(R) (GenomeDx Biosciences) to Predict Five-year Risk of Metastatic Prostate Cancer in Men Treated with Radical Prostatectomy. PLoS Curr 7. 10.1371/currents.eogt.761b81608129ed61b0b48d42c04f92a4.

Massey, J.C., Magagnoli, J., Sutton, S.S., Buckhaults, P.J., and Wyatt, M.D. (2024). Collateral damage of NUDT15 deficiency in cancer provides a cancer pharmacogenetic therapeutic window with thiopurines. bioRxiv. 10.1101/2024.04.08.588560.

Massie, C.E., Mills, I.G., and Lynch, A.G. (2017). The importance of DNA methylation in prostate cancer development. J Steroid Biochem Mol Biol 166, 1–15. 10.1016/j.jsbmb.2016.04.009.

Mateo, J., Carreira, S., Sandhu, S., Miranda, S., Mossop, H., Perez-Lopez, R., Nava Rodrigues, D., Robinson, D., Omlin, A., Tunariu, N., et al. (2015). DNA-Repair Defects and Olaparib in Metastatic Prostate Cancer. N Engl J Med 373, 1697–1708. 10.1056/NEJMoa1506859.

Mooijman, D., Dey, S.S., Boisset, J.C., Crosetto, N., and van Oudenaarden, A. (2016). Single-cell 5hmC sequencing reveals chromosome-wide cell-to-cell variability and enables lineage reconstruction. Nat Biotechnol 34, 852–856. 10.1038/nbt.3598.

NCI (2018). Cancer Stat Facts: Prostate Cancer. National Cancer Institute: Surveillance, Epidemiology, and End Results Program.

Nishiyama, A., and Nakanishi, M. (2021). Navigating the DNA methylation landscape of cancer. Trends Genet 37, 1012–1027. 10.1016/j.tig.2021.05.002.

Ongaba, T. (2023). ENOX1, CCDC122 AND LACC1 role in progression of prostate cancer. medRxiv, 2023.2010.2012.23296974. 10.1101/2023.10.12.23296974.

Picard (2019). Picard Toolkit. Broad Institute, GitHub repository. https://broadinstitute.github.io/picard/.

Piccolo, S.R., Sun, Y., Campbell, J.D., Lenburg, M.E., Bild, A.H., and Johnson, W.E. (2012). A single-sample microarray normalization method to facilitate personalized-medicine workflows. Genomics 100, 337–344. 10.1016/j.ygeno.2012.08.003.

Posa, I., Carvalho, S., Tavares, J., and Grosso, A.R. (2016). A pan-cancer analysis of MYC-PVT1 reveals CNV-unmediated deregulation and poor prognosis in renal carcinoma. Oncotarget 7, 47033–47041. 10.18632/oncotarget.9487.

Sarver, A.L., Murray, C.D., Temiz, N.A., Tseng, Y.Y., and Bagchi, A. (2016). MYC and PVT1 synergize to regulate RSPO1 levels in breast cancer. Cell Cycle 15, 881–885. 10.1080/15384101.2016.1149660.

Sharma, N.L., Massie, C.E., Ramos-Montoya, A., Zecchini, V., Scott, H.E., Lamb, A.D., MacArthur, S., Stark, R., Warren, A.Y., Mills, I.G., and Neal, D.E. (2013). The androgen receptor induces a distinct transcriptional program in castration-resistant prostate cancer in man. Cancer Cell 23, 35–47. 10.1016/j.ccr.2012.11.010.

Siegel, R.L., Giaquinto, A.N., and Jemal, A. (2024). Cancer statistics, 2024. CA: A Cancer Journal for Clinicians 74, 12–49. 10.3322/caac.21820.

Sjostrom, M., Zhao, S.G., Levy, S., Zhang, M., Ning, Y., Shrestha, R., Lundberg, A., Herberts, C., Foye, A., Aggarwal, R., et al. (2022). The 5-Hydroxymethylcytosine Landscape of Prostate Cancer. Cancer Res 82, 3888–3902. 10.1158/0008-5472.CAN-22-1123.

Song, C.X., Szulwach, K.E., Fu, Y., Dai, Q., Yi, C., Li, X., Li, Y., Chen, C.H., Zhang, W., Jian, X., et al. (2011). Selective chemical labeling reveals the genome-wide distribution of 5-hydroxymethylcytosine. Nat Biotechnol 29, 68–72. 10.1038/nbt.1732.

Steiner, T., Junker, K., Burkhardt, F., Braunsdorf, A., Janitzky, V., and Schubert, J. (2002). Gain in chromosome 8q correlates with early progression in hormonal treated prostate cancer. Eur Urol 41, 167–171. 10.1016/s0302-2838(01)00030-6.

Subramanian, A., Tamayo, P., Mootha, V.K., Mukherjee, S., Ebert, B.L., Gillette, M.A., Paulovich, A., Pomeroy, S.L., Golub, T.R., Lander, E.S., and Mesirov, J.P. (2005). Gene set enrichment analysis: a knowledge-based approach for interpreting genome-wide expression profiles. Proc Natl Acad Sci U S A 102, 15545–15550. 10.1073/pnas.0506580102.

Therneau, T.M. (2020). A Package for Survival Analysis in R [Internet]. Available from: https://CRAN.R-project.org/package=survival.

Tortolero-Luna, G., Zavala-Zegarra D., Pérez-Ríos N., Torres-Cintrón C.R., Ortiz-Ortiz K.J., Traverso-Ortiz M., Roman-Ruiz Y., Veguilla-Rosario I., Vázquez-Cubano N., Merced-Vélez M.F., Ojeda-Reyes G., Hayes-Vélez F.J., Ramos-Cordero M., López-Rodríguez A., Pérez-Rosa N. (2013). Cancer in Puerto Rico, 2006-2010: Incidence and Mortality. Puerto Rico Central Cancer Registry: San Juan, PR.

Tseng, Y.Y., and Bagchi, A. (2015). The PVT1-MYC duet in cancer. Mol Cell Oncol 2, e974467. 10.4161/23723556.2014.974467.

Tsui, K.H., Chiang, K.C., Lin, Y.H., Chang, K.S., Feng, T.H., and Juang, H.H. (2018). BTG2 is a tumor suppressor gene upregulated by p53 and PTEN in human bladder carcinoma cells. Cancer Med 7, 184–195. 10.1002/cam4.1263.

Vaarala, M.H., Hirvikoski, P., Kauppila, S., and Paavonen, T.K. (2012). Identification of androgen-regulated genes in human prostate. Mol Med Rep 6, 466–472. 10.3892/mmr.2012.956.

Wickham, H. (2016). ggplot2: Elegant Graphics for Data Analysis (Springer, New York, NY).

Wu, T., Hu, E., Xu, S., Chen, M., Guo, P., Dai, Z., Feng, T., Zhou, L., Tang, W., Zhan, L., et al. (2021). clusterProfiler 4.0: A universal enrichment tool for interpreting omics data. Innovation (Camb) 2, 100141. 10.1016/j.xinn.2021.100141.

Xiao, Z., Wu, W., Wu, C., Li, M., Sun, F., Zheng, L., Liu, G., Li, X., Yun, Z., Tang, J., et al. (2021). 5-Hydroxymethylcytosine signature in circulating cell-free DNA as a potential diagnostic factor for early-stage colorectal cancer and precancerous adenoma. Mol Oncol 15, 138–150. 10.1002/1878-0261.12833.

Xu, N., Wu, Y.P., Ke, Z.B., Liang, Y.C., Cai, H., Su, W.T., Tao, X., Chen, S.H., Zheng, Q.S., Wei, Y., and Xue, X.Y. (2019). Identification of key DNA methylation-driven genes in prostate adenocarcinoma: an integrative analysis of TCGA methylation data. J Transl Med 17, 311. 10.1186/s12967-019-2065-2.

Yan, Y., Zhou, B., Lee, Y.J., You, S., Freeman, M.R., and Yang, W. (2022). BoxCar and shotgun proteomic analyses reveal molecular networks regulated by UBR5 in prostate cancer. Proteomics 22, e2100172. 10.1002/pmic.202100172.

Yin, Z., Spitz, M.R., Babaian, R.J., Strom, S.S., Troncoso, P., and Kagan, J. (1999). Limiting the location of a putative human prostate cancer tumor suppressor gene at chromosome 13q14.3. Oncogene 18, 7576–7583. 10.1038/sj.onc.1203203.

Yokoyama, A., Kouketsu, T., Otsubo, Y., Noro, E., Sawatsubashi, S., Shima, H., Satoh, I., Kawamura, S., Suzuki, T., Igarashi, K., and Sugawara, A. (2021). Identification and Functional Characterization of a Novel Androgen Receptor Coregulator, EAP1. J Endocr Soc 5, bvab150. 10.1210/jendso/bvab150.

